# The timeout procedure in pediatric surgery - effective tool or lip service? A randomized prospective observational study

**DOI:** 10.1101/2020.10.15.20211466

**Authors:** Oliver J. Muensterer, Hendrik Kreutz, Alicia Poplawski, Jan Goedeke

## Abstract

**Background:** For over a decade, the preoperative timeout procedure has been implemented in most pediatric surgery units. In our hospital, a standardized team-timeout is performed before every operation. However, the impact of this intervention has not been systematically studied.

**Purpose:** This study evaluates whether purposefully-introduced errors during the timeout routine are picked up by the operating team members.

**Methods:** After ethics board approval and informed consent, deliberate errors were randomly and clandestinely introduced into the timeout routine for elective surgical procedures by a pediatric surgery attending. Errors were randomly selected among wrong name, site, side, allergy, intervention, birthdate, and gender items. The main outcome measure was how frequent an error was picked up by the team, and by whom.

**Results:** Over the course of 16 months, 1800 operations and timeouts were performed. Errors were randomly introduced in 120 cases (6.7%). Overall, 54% of the errors were picked up, the remainder went unnoticed. Errors were picked up most frequently by an anesthesiologists (64%), followed by nursing staff (28%), residents-in-training (6%) and medical students (1%).

**Conclusions:** Errors in the timeout routine go unnoticed by the team in almost half of cases. Therefore, even if preoperative timeout routines are strictly implemented, mistakes may be overlooked. Hence, the timeout procedure in its current form appears unreliable. Future developments may be useful to improve the quality of the surgical timeout and should be studied in detail.

## INTRODUCTION

In 2008 the World Health Organization (WHO) launched a Surgical Safety Checklist as part of the Global Patient Safety Challenge in an effort to reduce surgical morbidity and mortality worldwide.[1] Since then, the preoperative timeout procedure has been implemented in most operating rooms, and most pediatric surgery units. In our hospital, a standardized team-timeout routine, based on the WHO checklist, is performed before every operation.

Checklists are lists of crucial tasks to be addressed in a specific order so that no important steps are forgotten. They are often used in aerospace environments, where they have been extensively studied shown to improve aviation safety. [2] On this basis, surgical safety checklists were created to improve team communication, create a systematic and comprehensive review of critical datapoints, assure the execution of important tasks, and flatten the hierarchy that often characterizes the culture of surgical teams, [3] with an ultimate goal to improve patient safety. Checklists may be implemented in the form of mnemonics, printed lists, posters, or electronic means. The adaptability of checklists to the healthcare environment has been critically appraised, particularly because of the complexity of the individual items and their narrative, unreproducible measures of compliance, and variability in outcome.[4]

A systematic review of 20 published studies found that implementation of a surgical checklist had the potential of decreasing mortality by 47 to 62 percent, and morbidity by around one third.[5] Another review confirmed decreases in complications, mortality and surgical site infections by odds ratios (OR) of 0.59, 0.77, and 0.57, respectively.[6] Similarly, a large epidemiologic study demonstrated a reduction in mortality by an OR of 0.6.[7] In contemporary practice, over three quarters of hospitals worldwide use surgical checklist timeouts.[8]

On the other hand, a well-performed meta-analysis found mixed, less encouraging impact of surgical checklists on mortality and complications.[9] This report found substantial heterogeneity of the available studies on the topic, particularly regarding study design, setting, and surgical specialty. In a recent observational study, the interaction between the timeout participants was found to be complete in only half of 200 observed elective surgeries,[10] indicating that important items may be missed. This was confirmed in a study using snapshot audits that found team timeout errors in 40 to 60 percent of cases.[11]

Consequently, there remains considerable uncertainty regarding the quality and performance of the surgical checklist timeout procedure, including its true efficacy for picking up critical mistakes. [12] For checklists to be effective, however, it is not only important that the listed items be mentioned, but also that potential errors be picked up by the team. To date, there is practically no data on how often errors in the timeout procedure are effectively identified or overlooked. Therefore, this study evaluates whether purposefully-introduced random errors during the timeout routine are noticed by the team. This study was designed compliant with the CONSORT 2010 criteria. [13]

## METHODS

### Study design

Deliberate errors were randomly, intermittently, and clandestinely introduced into the surgical checklist timeout routine for elective pediatric surgical procedures by two attending pediatric surgeons. Errors were randomly selected among pre-defined categories including wrong name, intervention, site, side, allergy, birthdate, or gender using random lists and block sizes of 100 patients each. Randomization was performed by one of the authors. In order not to raise any suspicion by team members, fewer than 1 in 10 cases were selected for deliberate error placement, and cases were randomly selected from medical record numbers to avoid noticeable inclusion patterns.

Team members included the surgical resident or fellow, the anesthesiologist, anesthesia nursing, scrub nurse, as well as nursing and medical students. None of these team members were aware of the study.

### Timeout procedure

The standardized timeout strictly followed the format of the WHO Surgical Safety Checklist.[1] In brief, in our department, the attending pediatric surgical attending commences the timeout procedure by loudly verbalizing “timeout” in the operating room. A paper guide is available and can be viewed by the surgeon. At this time, all other non-essential activities must cease and the team’s attention focuses only on the timeout procedure. Using the written consent form of the patient, the surgeon verifies the patient’s name, birthdate, and the procedure to be performed, as well as the laterality if applicable. In interventions with laterality, the site is marked in the pre-surgical checklist before entering the operating suite. Special or potentially complicating issues are then mentioned. The surgeon checks that all imaging is displayed for the procedure, if applicable. Finally, the surgeon states whether preoperative antibiotic prophylaxis is necessary, whether there are allergies, or any critical steps anticipated, and if blood should be available for the operation. Subsequently, the anesthesiologist takes over by describing the type of anesthesia given, the patient’s weight, as well as any recorded allergies. The anesthesiologist confirms if antibiotic prophylaxis was given, describes number and types of intravascular access routes present, and whether blood products have been typed and crossed or are available in the operating room. Finally, the nursing staff confirms that the appropriate instrumentation for the procedure is available, sterile, and complete. They ask about the estimated time of the procedure and when the next patient should be called for, which is answered by the surgeon. The surgeon then asks if all team members agree with the issues noted during the timeout. Team members are instructed to verbalize any discrepancies. Unknown team members are asked to introduce themselves and their function. If all agree, the operation commences.

Until that point, there is a written policy in our department that no scalpels or cautery instruments are handed over, unless in emergency situations. The timeout is recorded on a standard surgical safety checklist paper form by the surgical resident.

### Error generation

Cases and type of error (name, intervention, site/side, allergy, gender, age) were selected for error introduction by random lists. The actual error was generated at the discretion of the attending surgeon. When applicable, typical first and surnames were substituted in lieu of the actual name, interventions were used that affected the same body region (for example, an orchidopexy instead of an inguinal hernia), sides were switched (right for left), allergies were declared when none were actually present, or vice versa. Gender was switched, and age was falsely modified within a 50% margin (a ten year old was falsely declared a child within a range of 5 to 15 years).

### Outcome parameters

The study’s main outcome measure focused on how frequent an error was picked up by the team, and by whom, as well as the type of error. Error detection was defined as someone in the operating room reacting to the error by verbal interjection by the end of the physician’s timeout. This was documented by a senior medical student or designee.

### Inclusion and exclusion criteria

Patients were included if they underwent elective, non-emergent surgery in our pediatric surgery operating room on our campus and the parents or legal guardian gave written consent for study participation. Exclusion criteria included refusal to participate by the parents, legal guardian or the patients themselves. Also, for safety reasons, patients were not included if they were operated in other operating rooms, facilities, or institutions.

### Ethical considerations

Ethics board approval was granted (approval number 837.105.17/10939, 2017). A particular request of the ethics board was that errors be corrected during the timeout procedure for safety reasons. Therefore, if the error was not positively identified by the end of the physician’s portion of the surgical safety checklist procedure, the attending pediatric surgeons verbalized a correction before proceeding, as illustrated in the following example:.

- Attending surgeon: “This is patient X, born on Y, who is a Z year old boy who is here for a right inguinal hernia repair. The site is marked and the consent has been signed by the parents. We are anticipating minor, negligible blood loss” (correct)
- Attending surgeon: “The patient is otherwise healthy and has no allergies” (incorrect - the patient has a documented allergy to amoxicillin)
- Anesthesia attending: “The patient is ASA class 1, we have an intravenous catheter in place in the right antecubital fossa, there is no blood crossed for this patient. Do you want any preoperative antibiotics?”
- Attending surgeon: “Preoperative antibiotics are not required in this clean case, thank you. But I just realized that the Mom told me that the patient does indeed have an allergy to amoxicillin. Please confirm.” (error corrected - entire team now knows about the amoxicillin allergy)
- Procedure continues as per protocol.

Informed consent by the parents or legal guardian of the patient was obtained. Patients 14 years of age or older were also asked to give their informed written consent for participation.

The only individuals informed about the study were the investigative team, the chief of anesthesia, as well as the director of operative services of the hospital. All agreed to observe absolute secrecy until study completion.

The study was registered at https://researchregistry.com (study number 2890).

### Statistics

From empirical evidence in our department, we assumed an error detection rate of over 90%. Our null-hypothesis was that less than 90% of errors would be discovered. Sample size was therefore calculated so that the lower end of the 95% Pearson confidence interval attained 90%. Actual error detection rate was estimated at 95% of errors. Clopper-Pearson confidence intervals (CI) were calculated for the detection rate. A pre-hoc analysis according to these presumptions calculated a minimum of 60 minor and 60 major errors necessary for adequate statistical power. However, we planned to continue the study until adequate sample size was reached. Detection rates were compared using the test of equal or given proportions. Post-hoc paired comparisons were calculated using the Benjamini-Hochberg procedure for multiple tests. The comparisons between team member roles were performed as a post-hoc analysis.

## RESULTS

Over the course of 16 months, a total of 1800 operations and timeouts were performed. Errors were randomly introduced in 120 of these cases (6.7%). In the 120 observed cases, in which errors were introduced, compliance with all elements of the timeout procedure was 100%. There were no instances in which a team member falsely corrected an unwarranted item. Also, no real, actual undetected errors were recorded during the study.

Overall, 54% of the errors were verbally challenged, the remainder went unnoticed. The total detection rate was 65 out of a total of 120 errors (54%). The detailed detection rates by types of error are found in table 1.

**Table 1:**
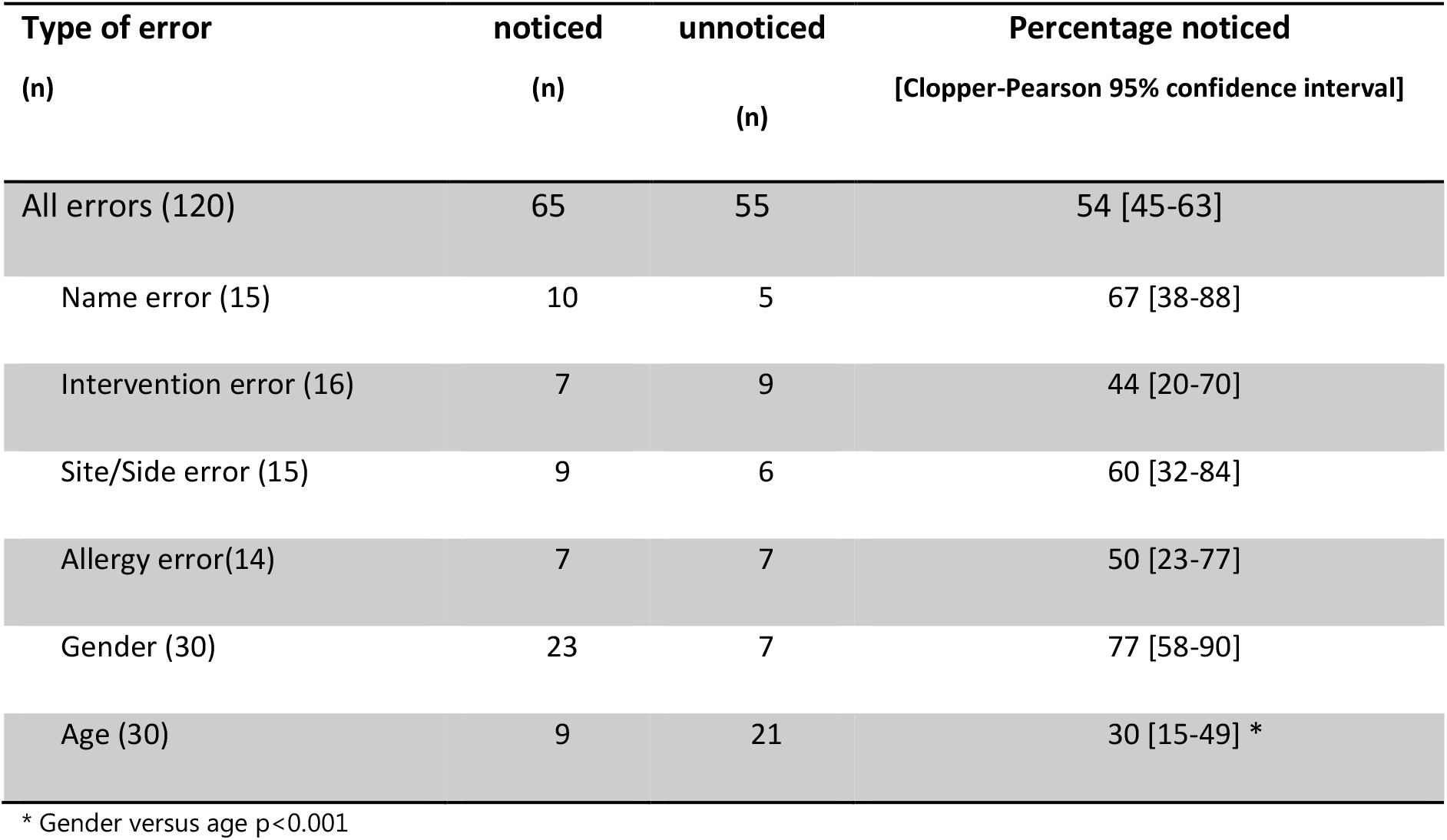
Error detection rates by type of error

The operative team was much more likely to identify a wrong gender mistake at a rate of 77% [95%CI 58-90%], compared to wrong age at a rate of 30% [95% CI 15-49%] (p<0.001).

Detection rates and their differences by profession or team function are graphically presented in figure 2. Errors were picked up most frequently by an anesthesiologists (64%), followed by nursing staff (28%), surgical residents or fellows in training (6%) and medical students (1%).

**Figure 1:**
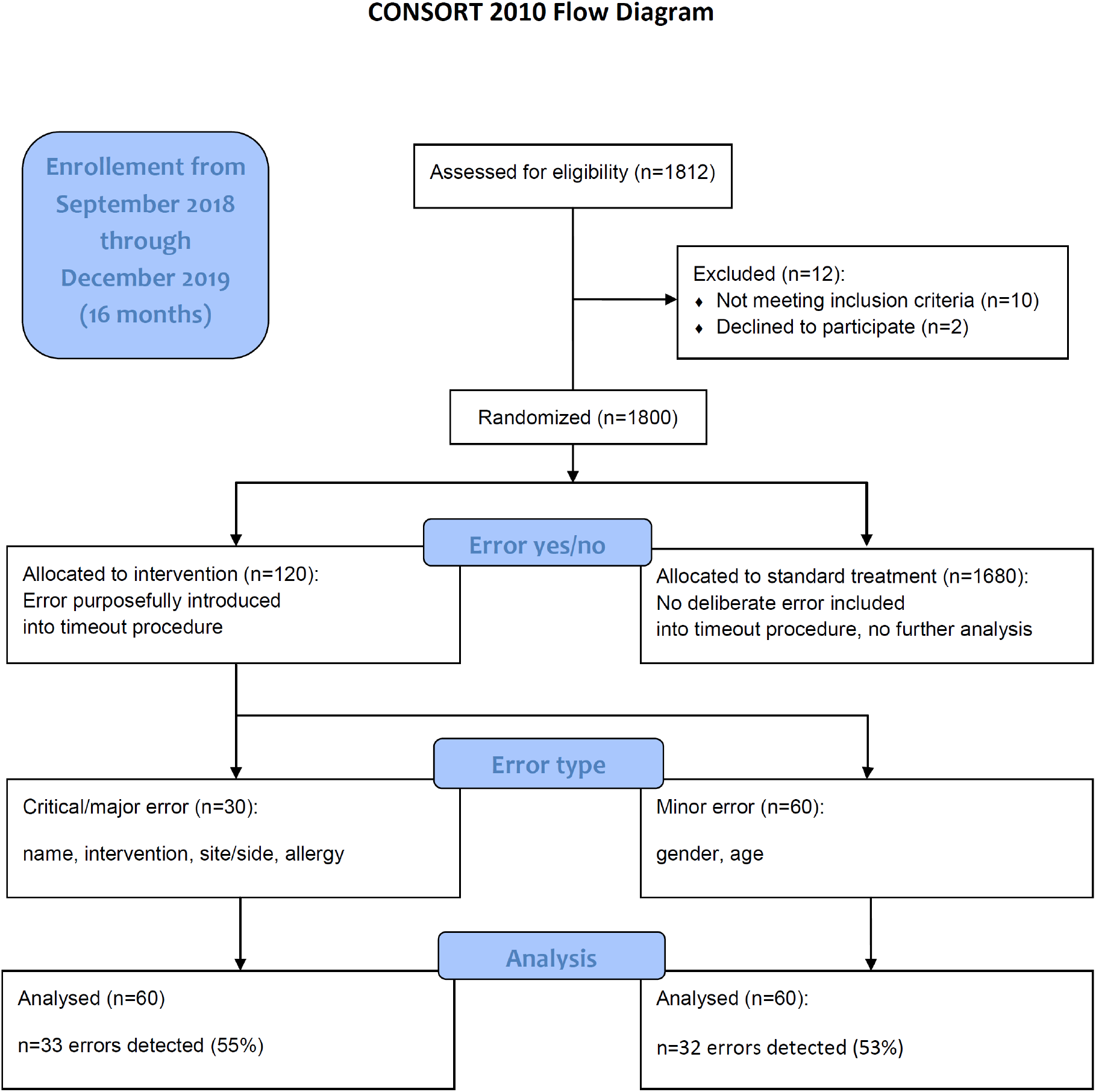
CONSORT 2010 Flowchart of the study participants.

**Figure 2:**
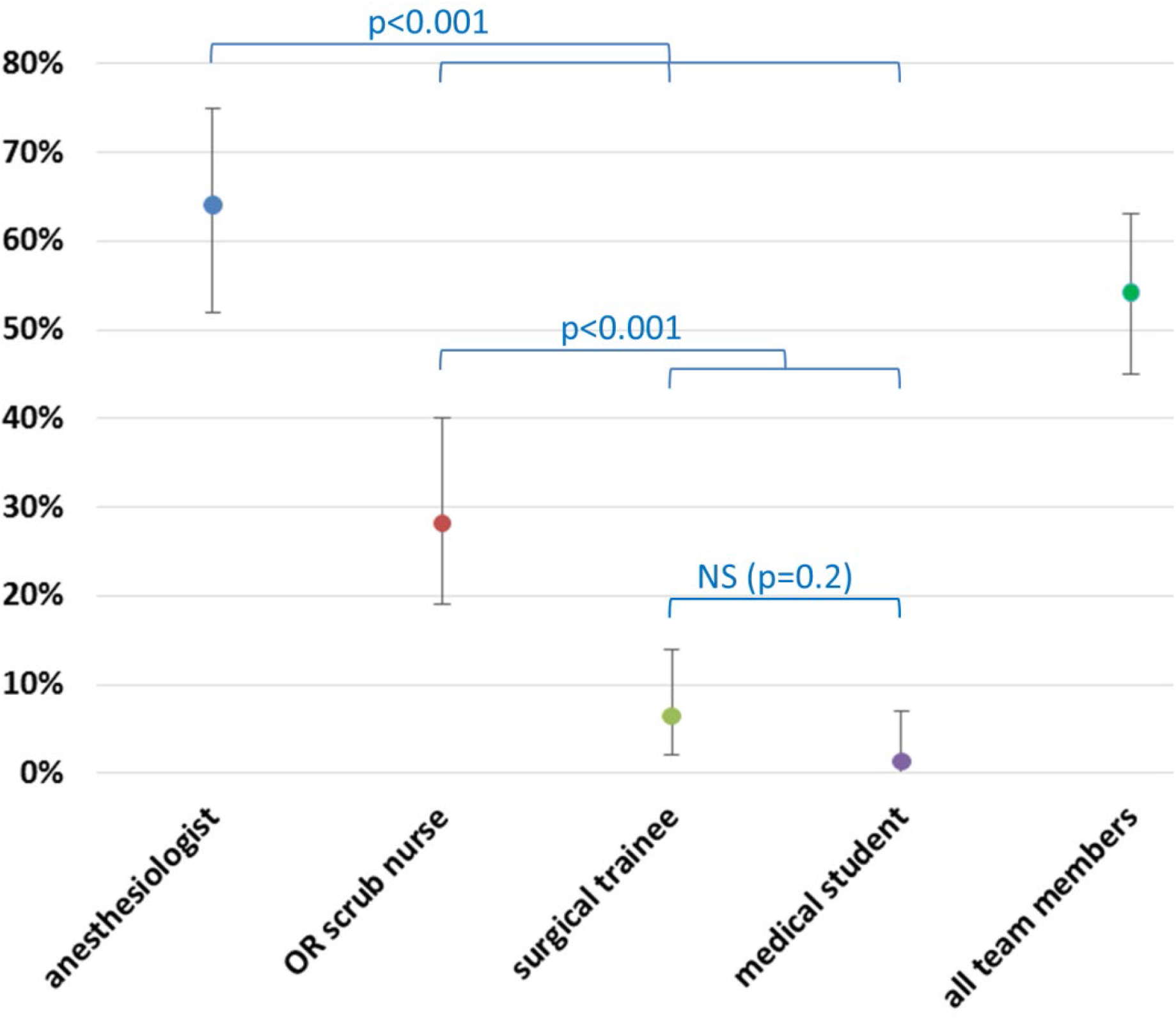
Error detection rates by profession/function in the operating team. Error bars represent 95% confidence intervals (NS - not significant).

## DISCUSSION

Since the introduction of the WHO Surgical Safety Checklist in 2008, it has been used globally in almost all countries in one form or another.[8] Despite before-after implementation studies showing a reduction in complications[12] and mortality,[7,14] significant barriers in timeout implementation have been identified.[15] Important factors seem to be cultural and procedural issues in specific healthcare settings.[16,17] Also, a study reported substantial deficiencies in medical school curricula regarding the teaching of surgical checklists and timeout procedures.[18] In contrast to the general trend, some authors have raised the concern that surgical timeout procedures may not reduce mortality after all.[9,19] Still others have found that wrong site and side surgery occurs despite the implementation of surgical timeout checklists.[20]

In children, the role of surgical checklist procedures in preventing morbidity and mortality is even less clear.[21] Studies show that there seems to be considerable variability in how timeout procedures are executed in pediatric surgery.[22,23] Also, the timeout procedure has been found to be incomplete im many cases, with important items omitted, forgotten, or left out.[24,25] One study from a tertiary pediatric surgery unit was able to document that the number of items discussed could be increased significantly from roughly half to nearly 100% by displaying laminated posters on the timeout procedure in the operating room.[26] A pre-post implementation study in Canada that included over 28,000 children showed no difference in complications after implementing a surgical safety checklist.[27] Attitudes may also play a role: In a recent survey, 94% of pediatric surgeons regularly used a surgical safety checklist, but only 63% would want it used on their own child, and only 55% thought that a timeout improved safety.[28]

There may be several putative factors decreasing the efficacy of surgical safety checklists. Stress and hastiness in the operating room may lead to omission of important data points. On the other hand, the timeout may be perceived as yet another bureaucratic burden imposed on busy clinicians by the hospital administration, rather than a helpful tool to bring the entire surgical team together onto one page at the beginning of the procedure.

Inattentiveness may be another factor that compromises the timeout. Before an operation, there are many issues to tackle, distracting individual team members, and keeping them from listening carefully. The surgical checklist timeout may thereby become a routine litany that is passively endured rather than actively participated in. Trainees, in particular, may not realize the importance of the timeout procedure, either because they have not been instructed appropriately, or because they are overwhelmed by other aspects of the operating room environment.

In this first experimental field study, we evaluated the attentiveness of the team during the timeout procedure in detecting randomly-sown errors. Particularly its undercover design allowed us to obtain more accurate information on how effective a timeout procedure truly is to pick up potentially hazardous errors. The higher detection rates of gender versus age may point to a baseline general attentiveness within the team that makes members react to obvious but not to subtle discrepancies. Therefore, the higher detection rate for gender may result from being the most salient feature of the patient addressed in the time out that is therefore most obvious to every member of the team. It is perhaps also the lowest risk to lower hierarchy team members if challenged.

In our study, anesthesiologists were much more likely than any other team member to pick up errors. Perhaps this is due to the relevance of their role in patient management, in which items such as age, weight and allergies are important. The higher detection rates may be related to anesthesia’s active role in the procedure itself. In our hospital, as in most institutions, both the anesthesiologist and the scrub nurse are required to actively verbalize certain parts of the checklist. Other reasons for higher anesthesiologist challenge rates may include their independence from surgical and other hospital workflow, as well as pertaining to a specialty that has a long history of patient safety recognition, training, and culture.[29]

Hierarchy may also play an important role in how likely a team member interjects when they notice a mistake. Modern safety culture and crew resource management calls for flat hierarchies to allow any member to verbalize discrepancies and potential hazards without fear of repercussion.[30]. A systematic review on the subject of authority and speaking up in the operating room show complex, multifactorial interactions.[31] In a simulation study, anesthesia trainees were more likely to challenge their attending’s decisions if they were more advanced in their training.[32] Others have shown negative hierarchical influence on the interactions and communication in a high-fidelity simulated operating room environment as well.[33] In our study, hierarchy was clearly related to detection rates. Trainees and students called out errors less often than attending anesthesiologists or professionally established scrub nurses.

The major weakness of this study is the single center design, which by nature can only describe our own local situation and circumstances. Other pediatric surgery teams may perform differently, despite the impetus by the WHO towards a standard protocol, which was the basis of the timeout procedure used in this study.. Also, the detection rates turned out to be much lower than initially presumed for the power analysis.

Another limitation is the early correction of verbalized errors by the attending at the end of the physician’s portion of the timeout procedure. Giving the team more time to correct an error may have increased the correction rates. However, the particular timing used in our study was a requirement set forth by the ethics committee to make sure that the risk of adverse consequences is minimized.

Unfortunately, the study design did not allow us to tease out the reasons for why an error was missed. We did not query the team members why they failed to point out an error. Future studies should survey the participants about inattentiveness, intimidation due to hierarchy, or other reasons.

The question remains how to improve timeout error detection rates. In a prospective field study, at least one member of the operating room team was actively distracted in more than one out of 10 procedures observed.[34] Similar deficiencies were recorded in a clandestine study in which medical students observed and recorded the timeout procedure in a large academic medical center.[35] In our opinion, attentiveness may perhaps be effectively facilitated by requiring interactive participation in the timeout by everybody in the room, including trainees and students. Although team members should regard themselves as a valuable part of the timeout safety check, systems safety science acknowledges that general imperatives such as “try harder” and “pay more attention” are very unlikely to be successful. Changing a deeply-seated cultural hierarchy requires a multimodal, long-term approach.

Checklists are one of the fields that lend themselves to computerized assistance. Modern data processing equipment could enhance the timeout procedure by providing an interactive, voice-controlled platform in which the team members sign in and then verbalize the items of a structured timeout procedure after being prompted. The system itself could check for plausibility according to the data in the electronic medical record, and at the same time verify issues such as patient weight, allergies, or relevant medications. It may also be programmed to check for completeness, for example whether all items and datapoints were mentioned. Mainthia et al have evaluated such an interactive electronic checklist system on an observational field study including 240 general surgery cases and found that compliance with addressing all core elements of the timeout procedure increased from about half of cases to 80 and 85 percent one and nine months after implementation of the system, respectively.[36] Similar improvements to compliance rates above 90 percent for all participating professions were found in another study after implementation of a surgical safety checklist integrated into the electronic medical record.[37] These improvements go beyond those recorded when simple posters are hung up in the operating room to improve safety checklist compliance.[38] Other interventions, such as implementation of a parent-centered script, did not increase checklist adherence.[39]

Finally, artificial intelligence combined with camera technology and motion detection could make sure that the patient is positively identified, and that the surgery is performed on the correct side or site. Future developments such as interactive, voice-activated platforms, computer-assisted timeout protocols, automatic patient identification, facial recognition and the use of artificial intelligence may potentially be useful to improve and enhance the quality of the surgical timeout procedure.

## CONCLUSIONS

In summary, almost half of deliberately sown errors in the timeout routine were not picked up by the team in this experimental field study. A wide variety of errors in the surgical timeout procedure were often not recognized. This may explain why the introduction of surgical safety checklists has had mixed impact on avoidance of errors in surgery. Factors associated with these findings may be inattentiveness, non-participation, false sense of hierarchical inferiority, or situational stress. This study indicates that the standard, structured verbal timeout procedure in its current form does not provide comprehensive protection from medical errors. Additional research is needed including evaluation of the potential benefits of advanced technology.

## Data Availability

Data are available from the authors upon reasonable request

## Patient consent for publication

Participants in this study or their guardians gave written consent for participation and publication of the resulting data in non-identifiable format.

## Data availability statement

The anonymized, non-identifiable data this study is based on is available upon request. Requests should be directed to the corresponding author. Reuse is permitted for scientific purposes and quality initiatives.

